# Accurate detection of 3D pelvic-floor-muscle strength for reforming clinical diagnosis and treatment of women’s pelvic floor dysfunction

**DOI:** 10.64898/2026.04.28.26351889

**Authors:** Tao Wang, Nannan Li, Hanbin Wang, Ying Zhou, Xiaojuan Yu, Qian Wang, Dongmei Wei, Rui Lian, Yi Luo, Xiaoyu Niu

**Affiliations:** Pelvic Floor Medical Center, Key Laboratory of Birth Defects and Related Diseases of Women and Children of Ministry of Education, West China Second University Hospital, Sichuan University, Chengdu, Sichuan,610041, China; Department of Obstetrics and Gynecology, Key Laboratory of Birth Defects and Related Diseases of Women and Children of Ministry of Education, West China Second University Hospital, Sichuan University, Chengdu, Sichuan, 610041, China; Micro/Nano Fabrication Laboratory, Microsystem & Terahertz Research Center, China Academy of Engineering Physics, Chengdu, 610200, China; Institute of Electronic Engineering, China Academy of Engineering Physics, Mianyang, 621900, China

**Author notes:** These authors contributed equally to this work (Tao Wang, Nannan Li).

## Abstract

Accurate and comprehensive evaluation of three-dimensional (3D) pelvic-floor-muscle (PFM) strength distributions are highly expected to play a crucial role in clinical early diagnosis and physiotherapy precise treatment of women’s pelvic-floor-dysfunction (PFD). However, clinically existing PFM-evaluating methods merely assess a rough and synthetical PFM strength from the whole vagina muscle tunnel, seriously restricting the development of PFD-related diagnostic methodologies and therapeutic interventions. Here, 3D complex female PFM-strength distributions have been accurately detected by developing a portable multi-channel PFM-pressure dynamic measuring system. Clinical trials demonstrate the superiority in high PFM-strength sensitivity and 3D spatial resolution, offering the opportunities to imply specific deficits in personalized PFM functions and customized interventions accordingly. Significantly, various subclinical PFM abnormalities can be identified by the 3D accurate PFM-strength distributions, which is not possible using traditional PFM-evaluating methods, providing a visual biomechanical foundation for clinical early diagnosis and physiotherapy precise treatment of PFD. Combined with the additional advance in physical comfortability, patients-friendliness universality, and stability without motion artifact achieved by the novel designed vaginal probe, this proof of concept research holds the promise for paradigm revolution in PFM pathological research, and promotes the transformation of clinical pelvic-floor medicine from empirical medicine to data-driven precision medicine.

## INTRODUCTION

Female pelvic floor muscle (PFM), which has the most complex anatomical structure among the numerous muscles of women’s body^1–4^, plays a crucial role in supporting pelvic organs.

Disruption of PFM integrity, which is generally induced by trauma secondary to childbirth, surgery, constipation, or chronic coughing, leads to weak PFM strength and even loss of PFM functions^5–8^. The pelvic floor dysfunction (PFD) with long course and high recurrence rate^9–11^, including pelvic organ prolapse (POP), urinary incontinence (UI), fecal incontinence, sexual dysfunction, and chronic pelvic pain, has a serious impact on the life quality of almost 25% of the female population^9^, and imposes substantial economic and public health burden^11–14^. Nowadays, PFD has been recognized as one of the five most prevalent chronic conditions that negatively impacts the quality of women’s life. Despite the serious burden of PFD, current diagnostic methodologies and therapeutic interventions are constrained by the inaccurate PFM assessment and limited understanding of the complex PFM architecture.

Clinically used PFM-evaluating methods, including vaginal digital palpation^15–17^, intravaginal force dynamometers^18–20^, intravaginal pressure perineometers^21,22^, and single-channel pelvic-floor electromyography (EMG)^23–25^, merely assess a rough and synthetical PFM strength of the whole vagina muscle tunnel, seriously restricting the diagnostic methodologies and therapeutic interventions of PFD-related disease. Vaginal digital palpation is the primary method to subjectively evaluate PFM strength in clinic according to a variety of grading scales such as Brink Scale^26^, Oxford Scale^17^, and Modified Oxford Scale^15,16^. Despite benefits from ease of operation, low cost, and high efficiency, the qualitative results highly dependent on doctors’ experience are prone to misdiagnosis. Although intravaginal force dynamometry or pressure perineometer are objective methods to evaluate PFM strength, they can only reflect the overall average strength of larger PFM groups during the tests. The lack of abilities in time-resolved dynamic testing and spatial-resolved location of multiple PFM directions limit their clinical application value. Nowadays, pelvic-floor EMG is widely used to dynamically monitoring a synthetical PFM strength of the whole vagina muscle by using commercially available intravaginal EMG probes with two stainless steel electrodes under professional guidance of physical therapists^27^. However, the electrical EMG signals usually exhibit poor reliability and validity due to the inevitable disturbance of motion artifact, low-frequency (LF) noise, pressure from peritoneal cavity, and crosstalk between muscles. Although EMG data is typically classified into Type I and Type II EMG, it still failed to characterize the activity signals of deep and specific small muscles surrounding vagina. Despite appearance of emerging 4-channel EMG technology^28^, the great challenges in the reliability and validity of electrical EMG signals still hinder their practical applications in clinic. Therefore, directly accurate detection of complex 3D PFM-strength distributions is urgently desired to promote clinical early diagnosis and precise physiotherapy treatment of PFD. This is of great significance for alleviating the medical burden of PFD population.

In this work, we demonstrated a direct method to accurately detect 3D PFM-strength distributions by developing a portable multi-channel PFM-pressure dynamic measuring system. The significant superiority in terms of high sensitivity with resolution of 1E-4 cmH_2_O and 3D PFM-strength distributions with 17-channel spatial resolution has been exhibited by comparing with traditional clinical PFM-evaluating methods. The high sensitivity with PFM-strength resolution of 1E-4 cmH_2_O permits observations of abnormally weak PFM strength state, allowing the identification of subclinical-state functional decay before clinical symptoms appearing. As a demonstration, subclinical PFM-strength abnormalities in the postpartum abdominal-wall-obesity (AWO) population have been identified, offering opportunities for early warning and primary prevention of PFD during the asymptomatic period. Significantly, the high PFM-strength resolution combined with 17-channel 3D spatial resolution allows for the identification of local PFM-strength defects, providing quantitative basis for the localization diagnosis of PFD. Clinical trials reveal that early PFD symptoms (e.g. unclosed vaginal orifice and stress urinary incontinence) could be linked with certain abnormal PFM-strength distribution characteristics, providing a visual biomechanical foundation for clinical early diagnosis and the classification diagnosis of PFD. In addition, the accurate 3D PFM-strength distributions could quantify local PFM-strength changes before and after treatments, providing objective indicators for curative effects. The design of individualized physical therapy programs would be expected. Combined with additional advance in physical comfortability, patients-friendliness universality, and stability without motion artifact offered by our novel designed vaginal probe, this work holds the promise for developing new diagnosis and exercise strategies for PFM pathological research, and has great potential for preclinical diagnosis and precise physiotherapy treatment of PFD. The clinical application of our method could significantly enhance diagnostic accuracy, treatment efficacy, and long-term management for PFD population, ultimately improving quality-of-life outcomes for this considerable patient population.

## RESULTS

### Hardware constitution

The portable multi-channel PFM-pressure dynamic measuring system is composed of a multi-contact semi-flexible pneumatized mace-shaped vaginal probe (PMVP) equipped with embedded gas pressure sensors (EGPSs), data acquisition processing and transmission modules, and terminal PC installed with self-developed analysis software (Fig. 1a). The PMVP consists of three parts: a dome-type contact for detecting abdominal pressure, multi-channel semi-flexible contacts to sense PFM-pressure distributions around vaginal wall, and the vaginal probe body with a handle (Fig. 1b). The dome-type contact is designed with hollow, flexible and round shape to allow the insertion into vaginal canal comfortably. The abdominal pressure could be propagated by the gas filled in the dome and then detected by the back-end EGPS. The multiple, hollow, and semi-flexible contacts with EGPSs are employed to dynamically monitoring multi-channel local PFM-strength distributions at various local areas of vaginal wall with different distance from fornix to vagina orifice simultaneously. The PMVP permits stable contact with vaginal wall, providing significant advance in physical comfortability, patients-friendliness universality, and stability without motion artifact.

**Fig. 1.**
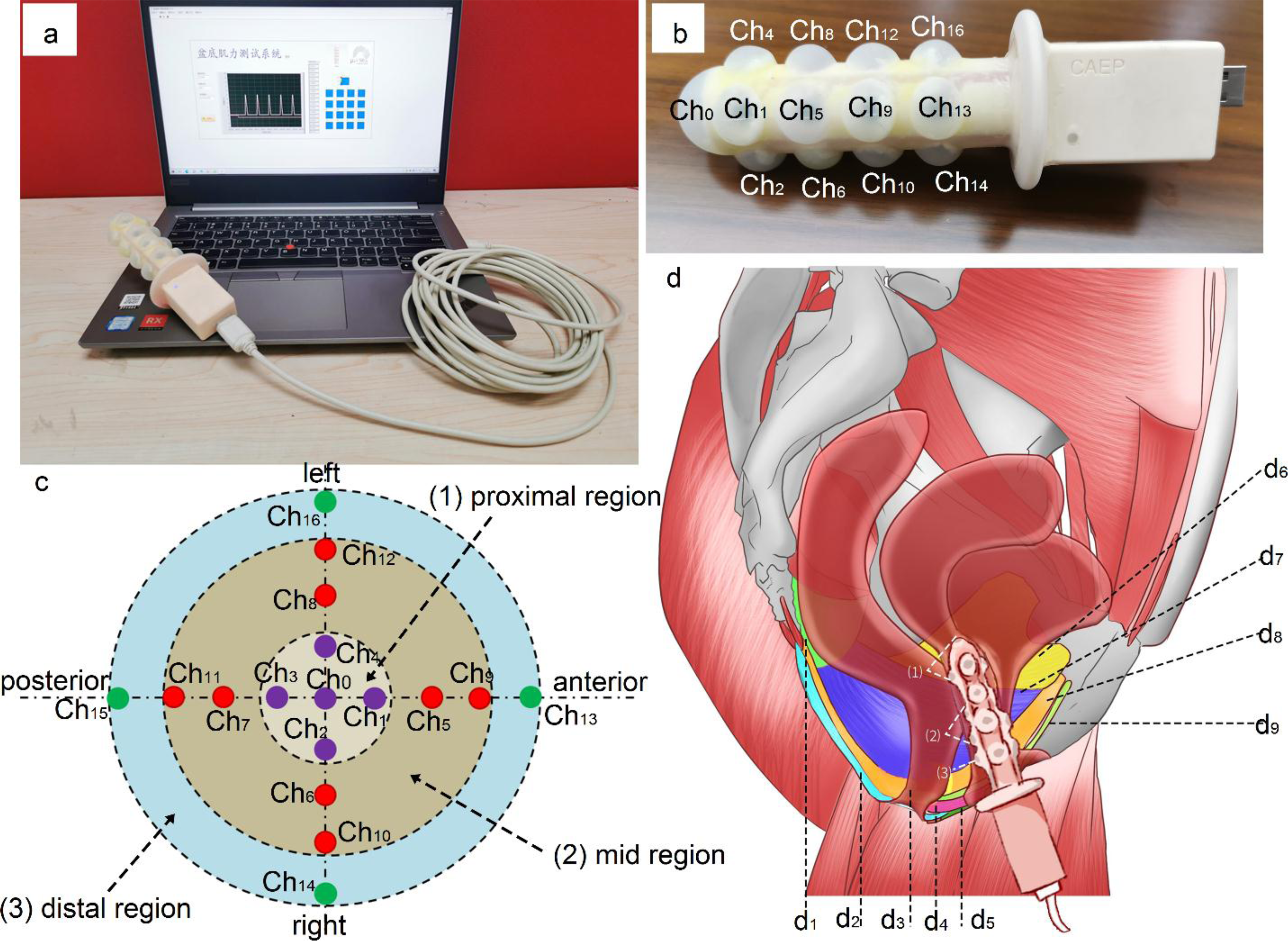
Schematic diagram of the direct method for accurately detecting 3D PFM-strength distributions. **a** The portable multi-channel PFM-pressure dynamic measuring system composed of a multi-contact semi-flexible pneumatized mace-shaped vaginal probe (PMVP), data acquisition processing and transmission modules, and terminal PC installed with self-developed analysis software. **b** The PMVP with 17-channel semi-flexible contacts equipped with embedded gas pressure sensors (EGPSs), labeling from CH0 to CH16. **c** Pelvic-floor distribution in a 2D format and the 17-channel contacts’ locations (dots) that are distributed according to the major PFM distributions in three regions: proximal (closer to the cervix), mid, and distal (closer to the introitus) regions. **d** Anatomical illustration of the PFM groups to be tested: d_1_−ischiococcygeus, d_2_−sphincter ani externus, d_3_−sphincter ani internus, d_4_−prerectal fibers, d_5_−deep transverse muscle of perineum, d_6_−obturator region, d_7_−lliococcygeus, d_8_−pubococcygeus, and d_9_−puborectalis.

As a demonstration, 17 semi-flexible contacts with EGPSs were fabricated on the surface of PMVP, which can collect 17-channel PFM-strength data for constructing 3D contour maps of PFM-strength distributions. To intuitively analyze PFM-strength distributions, the pelvic floor is distributed in a 2D format, and the multi-channel contact locations of PMVP (dots) are distributed according to the major muscle distributions in three regions: proximal (closer to the cervix), mid, and distal (closer to the introitus) regions (Fig. 1c). The 16 contacts are equally distributed into 4 rings with same pitch of 18 mm on the probe and the numbering rules are as follows: (1) four proximal-ring contacts close to fornix are equally located at anterior, right, posterior, and left vaginal wall in circumferential direction (CD), and labeled as CH1, CH2, CH3, and CH4 respectively; (2) eight mid-ring contacts (CH5 ∼ CH8; CH9 ∼ CH12) and four distal-ring contacts (CH13 ∼ CH16) are similarly distributed in three CDs on the probe; (3) the first contact of each ring, which is labeled as CH1, CH5, CH9, and CH13 respectively, distributes along the anterior vaginal wall in the longitudinal direction (LD) from cervix to introitus; (4) there is a domal contact labeled as CH0 along with four proximal-ring contacts to detect the abdominal pressure that transmits into proximal (closer to the cervix) region of vagina. The multi-channel-contact distributions on PMVP were designed to well match vagina structure and its rounding muscularis layer. The vagina is a fibromuscular tubular organ, which is comprised of internal mucosa, intermediate muscularis, and external adventitia layers, extending from the vulva to the cervix. The muscularis layer is composed of two layers of smooth muscle cells (SMCs): an inner layer with circumferentially oriented SMCs and an outer layer with longitudinally oriented SMCs^29^. The PFM-detecting contacts distributed on the surface of PMVP is consistent with the direction of the two layers SMCs. The multi-channel contacts are employed to detect PFM groups at different locations, including ischiococcygeus, sphincter ani externus, sphincter ani internus, prerectal fibers, deep transverse muscle of perineum, obturator region, lliococcygeus, pubococcygeus, and puborectalis (Fig. 1d). This fixed-point numbering design provides a reliable diagnostic basis for the precise location and analysis of abnormal PFM strength. The data acquisition processing and transmission system was miniaturized and integrated into the probe handle, significantly enhancing the portability of the measurement system. A self-developed analysis software installed in terminal PC was used to record and analyze the multi-channel PFM-strength data. The PMVP with high-precision miniaturized EGPSs exhibits real-time multi-channel dynamic detecting capability with high accuracy resolution of 1E-4 cmH_2_O. Finally, 3D contour maps of PFM-strength distributions can be constructed by using the measured multi-channel PFM-strength curvatures at different locations, facilitating to access the complex 3D PFM architectures and functions and then provide direct and visible evidences for clinical diagnosis and pathological analysis.

Compared with currently existing PFM-strength evaluating equipment, our portable multi-channel PFM-pressure dynamic measuring system exhibits significant advantages as outlined in Table 1. The PMVP design addressed the fitting problem between probe and vagina for various subjects and even for subjects experiencing vaginal laxity. The flexible gas-filled contacts can change their volumes according to users’ physiological conditions, permitting PMVP in vagina firmly without disturbance of motion artifact, and enabling patients-friendliness universality by balancing comfort during test process and accuracy of the measured data. The PMVP equipped with high-precision miniaturized EGPSs exhibits real-time multi-channel dynamic PFM-pressure detecting capability with high accuracy resolution of 1E-4 cmH_2_O. Compared with the clinically used pelvic-floor EMG signals, our multi-channel PFM-strength data exhibits excellent reliability and validity without disturbance from low-frequency (LF) noise, crosstalk between muscles, and abdominal pressure. Significantly, abdominal pressure that generally confused in vaginal cavity has been separately detected by using the EGPSs at the dome and in the neighboring proximal region. Accurate 3D mapping of PFM-strength distribution can be achieved based on the measured multi-channel PFM strength data by using our self-developed analysis software. Furthermore, the complex post-processing equipment and the physician experience or professional guidance has also been released, expanding future potential applications of this method in various individualized diagnosis and treatment of PFD. Clinically, the portable multi-channel PFM-pressure dynamic measuring system can be quickly deployed in the outpatient department, operating room, or even community settings, enhancing the operability of PFD-related screening.

**Table 1.** Features of our portable multi-channel PFM measurement system.

|  | **Hardware composition** | **Bottlenecks to be addressed** | **Advantages or innovations** |
| --- | --- | --- | --- |
| 1 | PMVP | ● Stability problem induced by probe slipping from vagina<br>● Difficult balance between fitment and comfort<br>● Unattainable universality for various subjects | ● Stability without motion artifact<br>● Comfortability<br>● Universality for various subjects |
| 2 | Multi-channel miniaturized EGPSs | ● Rough and synthetical PFM-strength value<br>● Poor reliability and validity<br>● Distinguish PFM strength from different muscle groups | ● High sensitivity of 1E-4 cmH<sub>2</sub>O<br>● High reliability and validity<br>● 17-channel PFM strength from different muscles |
| 3 | A dome contact with neighboring inner-ring contacts | ● PFM pressure confused by abdominal pressure | ● Abdominal pressure can be separately measured |
| 4 | Terminal PC installed with self-developed analysis software | ● Complex post-processing equipment required<br>● Dependent on physician experience or professional guidance<br>● Limited clinical utility | ● Ease of operation<br>● Independent on physician experience or professional guidance<br>● Portability for mobile applications |

### Clinical verification

To preliminarily demonstrate the feasibility and superiority of our PFM-strength measuring method, clinical PFM strength acquisition comparisons were conducted using our novel multi-channel PFM-strength measuring method and commercial pelvic-floor EMG method (MLD B4Plus, Nanjing Mai Land Medical Technology Co., Ltd., China). Participants included 29 healthy women without any PFD symptoms and without postpartum restriction. The Glazer protocols were used as the standard assessment actions, consisting of pre-rest baseline test, five rapid contractions, and five tonic contractions. Details of the standard Glazer protocol procedure are shown in the Materials and Methods Section. The rapid contraction can be used to assess functions of the fast-twitch muscle fibers, while the tonic contraction can be used to assess functions of the slow-twitch muscle fibers. The typical EMG signals and the corresponding multi-channel PFM-strength distributions are shown in Fig. 2a and Fig. 2b, respectively. The similar rapid contraction and tonic contraction trends, which reveals functions of fast-twitch and slow-twitch muscle fibers, preliminarily exhibit the feasibility of our novel PFM-strength measurement system. In addition, comparisons of clinical PFM-strength results for typical healthy and unhealthy subjects has also been conducted by using commonly pelvic-floor EMG method and our novel method respectively. As shown in Supplementary Fig. 1, the consistency with clinical conclusion further confirmed the validity of our PFM-strength measuring method.

**Fig. 2.**
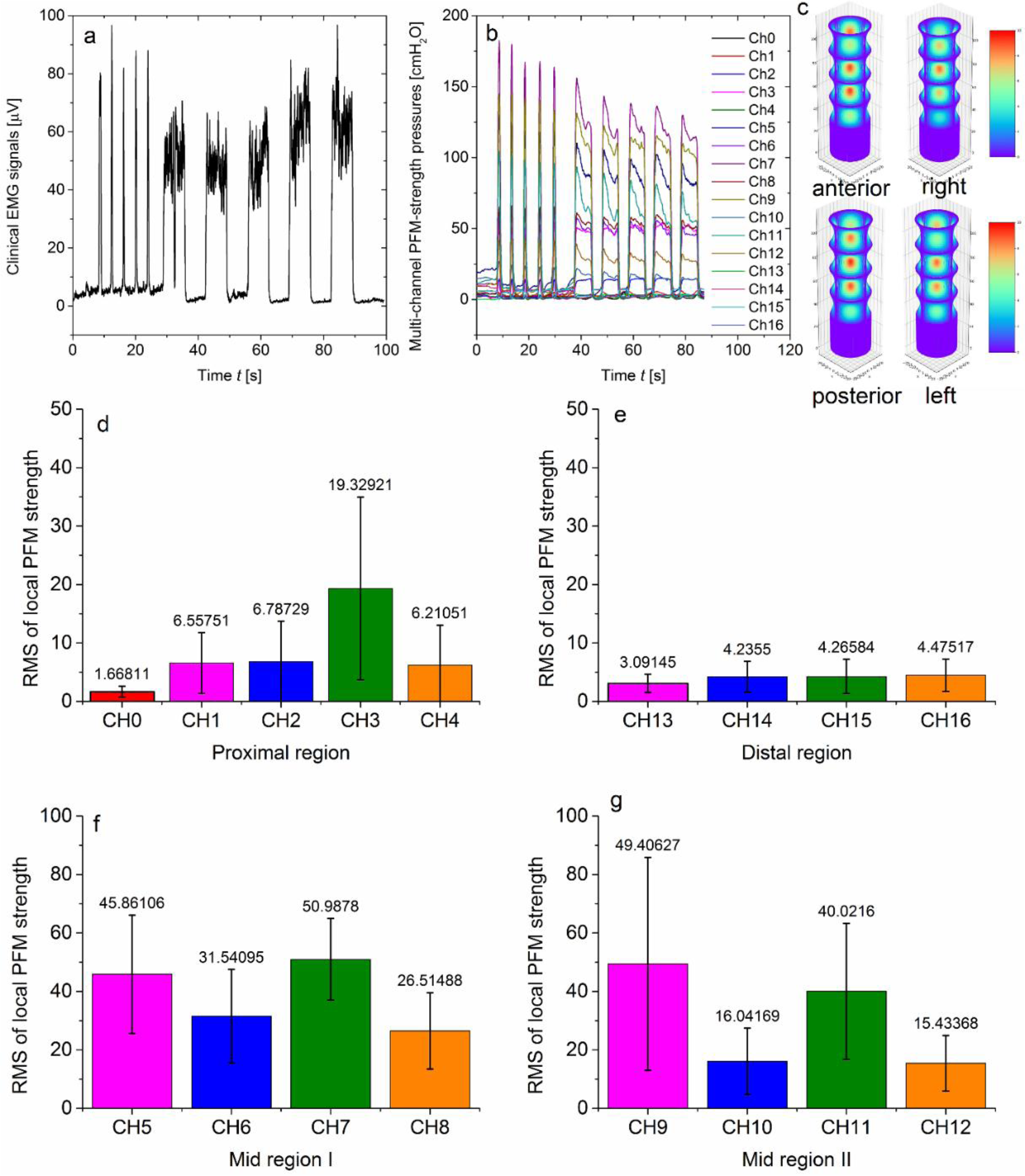
Demonstration of feasibility and superiority of our accurate 3D PFM-strength distributions measuring method. **a** Typical EMG signals of a healthy subject by using commercial pelvic-floor EMG method. **b** The corresponding multi-channel PFM-strength distributions for the same subject in **a**. The similar rapid contraction and tonic contraction trends, which reveals functions of fast-twitch and slow-twitch muscle fibers, preliminarily exhibit the feasibility of our novel PFM-strength measurement system. **c** The 3D PFM-strength distributions in four directions, which have been constructed by using our measured multi-channel PFM pressures shown in **b**. **d** The statistical RMS of PFM strengths at each local channel (CH0 ∼ CH4) in the proximal region. **e** The statistical RMS of PFM strengths at each local channel (CH13 ∼ CH16) in the distal region. **f** The statistical RMS of PFM strengths at each local channel (CH5 ∼ CH8) in the mid region I. **g** The statistical RMS of PFM strengths at each local channel (CH9 ∼ CH12) in the mid region II.

Significantly, our multi-channel PFM-strength measuring results could provide more valuable PFM-strength information than the commonly used pelvic-floor EMG method, which is of great importance for clinical early diagnosis and precise physiotherapy treatment of PFD. The 3D PFM-strength distributions in LD and CD directions of the entire pelvic floor have been constructed by using our measured multi-channel PFM pressures (Fig. 2c), which is well agreement with the two SMCs layer distributions: the inner circumferentially oriented SMCs layer and the outer longitudinally oriented SMCs layer^29^. This is helpful for doctors to quickly identify the weak PFM locations. The entire pelvic floor was divided into three regions: the proximal, midpiece, and distal regions. The multi-channel PFM-strength data in each region for all the 29 healthy participants are summarized in Figs. 2d−2g, respectively. The PFM-strength curves in each channel were converted to RMS values. The RMS of PFM strength at each channel location in four LD directions was indicated by different color bands for comparison. The PFM strength in the proximal region of vagina could be evaluated by the domal channel (CH0) and the neighbouring 4 channels in proximal-ring contacts (from CH11 to CH4), as shown in Fig. 2d. There are significant differences between CH3 and other channels (*p* < 0.001). The PFM strength distribution in the proximal region can be utilized to access the abdominal pressure that propagated into vaginal cavity, exhibiting the highest pressure point at the posterior fornix with good coherence with anatomy and physiology. The RMS of PFM-strength distribution in the distal region of vagina could be evaluated by the 4 symmetrical channels in distal-ring contacts (from CH13 to CH16), as shown in Fig. 2e. There is no significant statistical difference between the four distal channels (*p* = 0.18), revealing the mechanical supporting ability of the vaginal orifice offered by the richly innervated sphincter structure of circumferentially aligned SMCs^30,31^. The relatively low pressure agrees with the fact of lacking strong muscles at the orifice of vagina. The pathogenesis of unclosed vaginal orifice could be revealed, assisting to find the risky factor or the accompanied feature of stress urinary incontinence. Compared the PFM distribution in proximal and distal regions, the longstanding disagreement whether the proximal vagina has equal^32^ or greater^33,34^ relative smooth muscle content than the distal vagina can be addressed. In this way, accurate 3D PFM-strength distributions in the proximal and distal region, which are generally neglected in traditional PFM-evaluating methods, provide great significance for paradigm revolution in PFM pathological research and revealing new pathology of PFD.

The midpiece region of PFM could be evaluated by 8 symmetrical channels (from CH5 to Ch12), as shown in Figs. 2f and 2g, exhibiting the main force bearing role of the pelvic organs. As consistent with the cognitive conclusion of clinicians, the midpiece region always displays much stronger PFM strength than the proximal and distal regions. Compared with the synthetical PFM strength of the whole vagina muscle without distinguishing different muscle groups in traditional PFM-evaluating methods, 8-channel local PFM-strength distributions with significant differences (*p* < 0.001) in the mid region have been found for the 29 healthy women. It has been found that local PFM strength from puborectalis muscle on the left and right sides of vaginal wall is generally lower than 80 cmH_2_O. The maximum local PFM strength in the mid-I region and in the mid-II region as shown in Figs. 2f and 2g, which are generally high than 100 cmH_2_O, are always in two diagonally opposite directions of the anterior and posterior vaginal wall of midpiece region. This method reveals that it is not the bilateral puborectalis muscles on the left and right sides of vaginal wall but the anal circumference muscles of the pelvic cavity plays a dominant role during vaginal contraction. This is consistent with the PFM distribution characteristics, where the posterior pelvic perirectal muscles are the most significant proportion of PFM. Specifically, the strongest local PFM strengths are always from CH7 and CH9, which is consistent with the oblique line of levator ani muscle. The multi-channel PFM-strength distribution in the mid region is consistent with the anatomic differences on the contractile properties of the vagina, which have not been found in traditional PFM-evaluating methods^15–25^. These observations should be attributed to the significant superiority of our novel PFM detecting method in terms of high sensitivity with resolution of 1E-4 cmH_2_O and 3D PFM-strength distributions with 17-channel spatial resolution. For different healthy women, human-to-human variability in PFM-strength distributions have been exhibited. Specifically, the individual difference in PFM-strength distributions offer the opportunities for precisely customizing personalized treatment strategies to prevent PFD occurrence.

### PFM-strength distributions for postpartum women with different symptoms

Accurate 3D PFM-strength distributions play an important role in analyzing various disease subtypes, facilitating to clinical early diagnosis and precise physiotherapy treatment of PFD. Comprehensive multi-channel PFM-strength measurements have been performed on multiple sub-groups with various symptoms including abdominal wall obesity (AWO), unclosed vaginal orifice (UVO), and stress urinary incontinence (SUI). Symptomatic and intractable PFD diseases are always developed from alterations and damage to the PFM structures which is related to above preclinical or clinical symptoms with abnormal PFM-strength distributions after pregnancy and delivery. It has been demonstrated that different subtypes presented distinct mechanical markers within the PFM-strength distribution characteristics. Beyond the ability of currently reported PFM-strength assessing methods^15–25^, our novel PFM detecting method could successfully link typical PFD-related symptoms with certain PFM-strength-distribution characteristics. This discovery provides a visual biomechanical foundation for early diagnosis and classification diagnosis of PFD. Moreover, this research establishes an accurate 3D parameter framework for target localization and treatment planning within individualized physical therapy programs.

#### (1) PFM-strength characteristics of postpartum AWO subgroup

The high PFM-strength resolution of our method allows the identification of subclinical-state functional decay before clinical symptoms appearing. Slight changes in PFM strength often precede the appearance of clinical symptoms. However, such early change is generally far below the detection threshold of traditional PFM-evaluation methods in clinic. As a demonstration, subclinical PFM-strength abnormalities in 22 postpartum AWO subjects have been idnetified by using our accurate 3D PFM-strength detecting method. A comprehensive pelvic floor examination including sequential vaginal digital palpation, intravaginal EMG, and our real-time multi-channel dynamic PFM pressure measuring method has been performed for the postpartum AWO women. To exclude the influence of delivery mode on the 3D PFM-strength distributions, half of the postpartum AWO subjects had experienced cesarean section (CS) and half of them had experienced vaginal delivery (VD). In this work, the clinical EMG outcomes of the postpartum subjects with AWO ranged from 55.1 to 81.1, demonstrating that EMG scores cannot reflect AWO effects. By contrast, comprehensive 3D PFM-strength distributions provide specific characteristics for the subclinical AWO symptoms (Fig. 3).

**Fig. 3.**
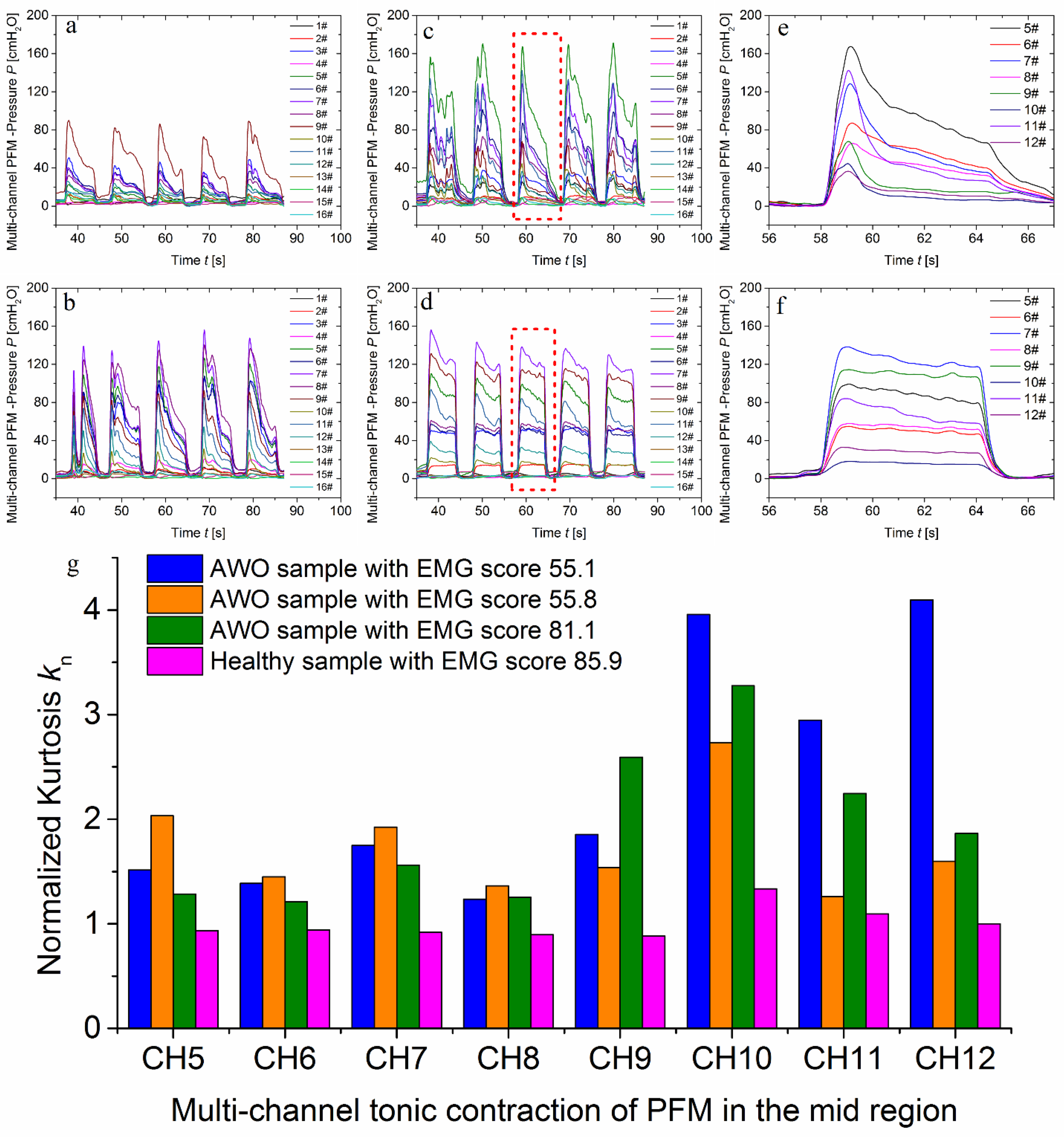
Comparison of PFM-strength characteristics for typical postpartum AWO subjects and a healthy subject. **a** Multi-channel PFM-strength distribution for a postpartum AWO subject whose EMG score was 55.8. **b** Multi-channel PFM-strength distribution for a postpartum AWO suject whose EMG score was 55.1. **c** Multi-channel PFM-strength distribution for a postpartum AWO subject whose EMG score was 81.1. **d** Multi-channel PFM-strength distribution for a healthy subject whose EMG score was 85.9. **e** The local magnified view of PFM-strength distributions in the midpiece region within the dashed box in **c**. **f** The local magnified view of PFM-strength distributions in the midpiece region within the dashed box in **d**. **g** The normalized Kurtosis comparison for the multi-channel tonic contraction of PFM in the mid region.

Compared with the measured multi-channel PFM-pressure of postpartum AWO subjects (Fig. 3a-c) with that of healthy subjects (Fig. 3d), a notable descending trend in each-channel PFM pressure values during tonic contraction was observed (Fig. 3e). This is mainly due to the difficulty of maintaining PFM strength during the tonic contraction phase for AWO subjects who have a decreased muscle ratio and poor muscle-strength endurance. This finding suggests a correlation between AWO and abnormal PFM function with a spiky activity pattern in slow muscle fibers (Fig. 3e), thereby further revealing why women with AWO are prone to PFD. Significantly, this also reveals the accuracy of our PFM detecting method, which is completely in line with clinical practice.

To quantitatively compare the PFM curve morphology during tonic contraction of the AWO subgroup (Fig. 3e) and that of the healthy group (Fig. 3f), kurtosis coefficient was introduced:

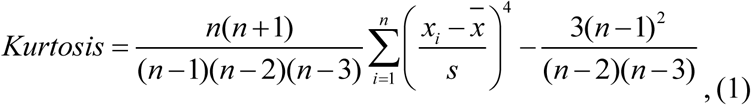

where *s* is the standard deviation of the samples on each channel curve as shown in Figs. 3a−3d, *n* is the sample size, and *x̄* is the mean of the samples. For ease of comparison, the kurtosis coefficient of multi-channel PFM-strength in the mid region during tonic contraction was normalized for a typical healthy subject. Figure 3g shows the comparison of normalized Kurtosis *k*_n_ of multi-channel PFM-strength in the mid region during tonic contraction for three typical AWO subjects and the typical healthy subject. A much higher *k*_n_, representing stronger steepness of the spiky activity pattern in slow muscle fibers, has been observed for each channel of the PFM-strength distributions in the mid region of AWO subjects. The notable descending trend in PFM strength during tonic contractions with high Kurtosis value was observed for many postpartum AWO subjects, attributing to the high PFM-strength resolution of our method. Specifically, both pelvic floor muscles and abdominal wall muscles collaboratively contribute to the maintenance of body core musculature stability. Dysfunction in PFM may lead to the core musculature instability, adversely affecting the normal contraction and support capabilities of the abdominal wall muscles^35^. Furthermore, we have distinctly identified the functional characteristics of slow-twitch muscle fibers (Type I muscle fibers), which are essential for sustained relatively long-time muscle contraction and postural maintenance. If there is dysfunction within these slow-twitch fibers, it could result in diminished support capacity for these muscles, thereby impacting coordination and stability within the abdominal wall musculature^36^. In relation to weight gain and changes in abdominal pressure during pregnancy, it is important to note that weight gain places an additional burden on the pelvic floor muscles. Throughout pregnancy, a woman’s body experiences significant weight increases due to fetal growth and fat accumulation. This added weight elevates abdominal pressure, imposing greater strain on PFM and potentially leading to dysfunction during long-time muscle contraction^36^. Simultaneously, fluctuations in abdominal pressure also affect pelvic floor functionality. As gestation progresses and uterine size expands, alterations occur in a pregnant woman’s abdominal pressure. Such variations can disrupt normal PFM functions, resulting in decreased contraction capacity shown as the spiky pattern in multi-channel PFM-pressure curves during tonic and sustained contraction. The observation of notable descending trend in PFM strength during tonic contractions with high Kurtosis value may provide one of biomarkers for early intervention of PFD. In this way, the high PFM-strength resolution of our method allows the identification of subclinical-state functional decay before clinical symptoms appear, offering opportunities for early warning and primary prevention and even for breaking through the dilemma of missed diagnosis during the asymptomatic period.

#### (2) PFM-strength characteristic of postpartum UVO subgroup

For 15 postpartum subjects with UVO symptoms, more than 85% of them had experienced VD. VD assisted by forceps, vacuum extraction, and even episiotomy tends to directly damage the muscles at the orifice of the vagina, which is the main reason of pathological process of UVO. Although UVO symptoms are generally believed to be related to an early stage of SUI, there is still a lack of effective methods to conveniently characterize the degree of UVO in clinic. In the distal region close to the introitus, SMCs appear to be scattered and unorganized^30,37^, leading to weak contractile forces difficult to be detected. The UVO degree is roughly judged by means of vaginal digital palpation and then further diagnosis by ultrasound in clinic nowadays. No accurate and convenient measurement method is available in clinic for evaluating the status of UVO. In this work, for the 15 postpartum subjects with UVO symptoms, the clinical EMG outcomes, whose score ranged from 10.1 to 82.5, cannot be used to assess the state and degree of UVO. By contrast, our multi-channel PFM-strength distributions in the distal region could quantitatively reveal the UVO degree. In particular, the 4-channel PFM-strength distributions in the distal region in rapid contraction and tonic contraction trends could reflect the symptoms of UVO. The multi-channel PFM-strength-distribution in the distal region of healthy subjects has been compared with that of postpartum UVO subjects (Fig. 4). Figure 4a shows the comparison of RMS of local PFM strength at each channel in the distal region for three typical postpartum UVO subjects with different EMG scores and a typical healthy subject. Much lower RMS value of local PFM strength at each channel in the distal region has been found for postpartum UVO subjects. The corresponding multi-channel PFM-strength distributions in the distal region are shown in Supplementary Fig. 2. It can be found that the PFM pressure in the distal region of UVO subjects are always lower than 10 cmH_2_O (Fig. 4b and Supplementary Figure 2a−c). For healthy subject, the maximum PFM pressure of at least two channels is over 30 cmH_2_O at rapid contraction and over 20 cmH_2_O at tonic contraction in the distal region (Fig. 4c). The weakness degree of amplitude of PFM strength in the distal region could reflect UVO degree, which has been confirmed by the comparison of the ultrasound results shown in the insets of Fig. 4b and c respectively. The capability to assess UVO degrees by monitoring the weak PFM-strength distributions in the distal region should be attributed to combination of high PFM-strength resolution of 1E-4 cmH_2_O and 3D PFM-strength distributions with 17-channel spatial resolution of our PFM-strength detecting method. This work provides an indirect diagnosis for judging UVO which is also one of the main causes of SUI.

**Fig. 4.**
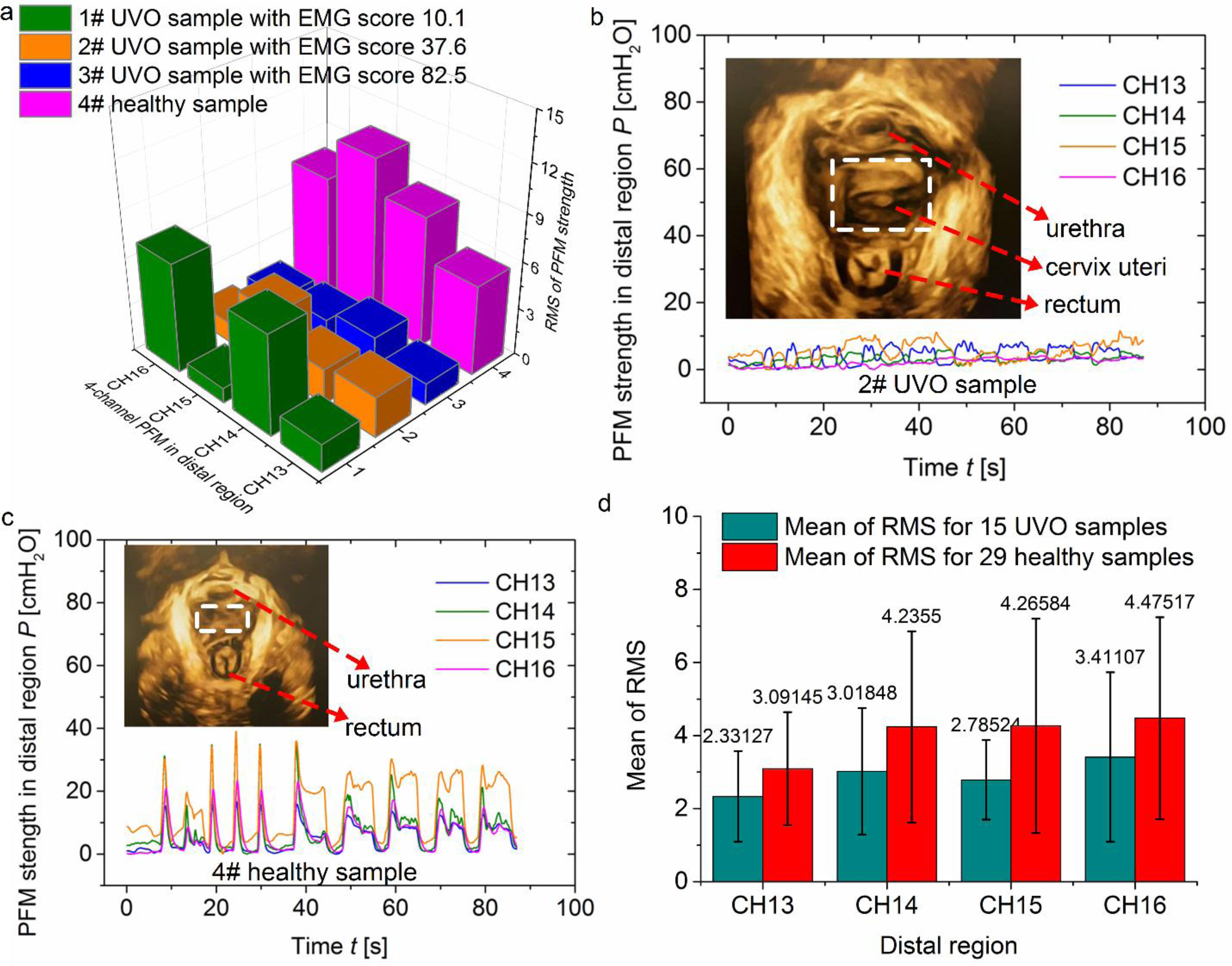
Comparison of PFM-strength characteristics in distal region for postpartum UVO subgroup and healthy group. **a** The comparison of RMS of local PFM strength at each channel in the distal region for three typical postpartum UVO subjects with different EMG scores and a typical healthy subject. **b** Four-channel PFM-strength distributions in the distal region of 2**#** UVO subject in **a**. **c** Four-channel PFM-strength distributions in the distal region of 4**#** healthy subject in **a**. **d** The statistical RMS of PFM strengths at each local channel (CH13 ∼ CH16) in the distal region for 15 postpartum UVO subjects and 29 healthy subjects.

To quantitatively reveal the characteristics of PFM-strength distributions for UVO subgroup, the statistical RMS of PFM strengths at each local channel (CH13 ∼ CH16) in the distal region have been analyzed for 15 postpartum UVO subjects and 29 healthy subjects (Fig. 4d). Much lower statistical RMS value of local PFM strength at each channel in the distal region has been found for postpartum UVO subgroup. The primary pathological mechanism underlying incomplete closure of the vaginal orifice is attributed to the disruption of the supportive function provided by PFM groups, such as the pubococcygeus and puborectalis muscles. These muscles constitute a dynamic structure essential for maintaining vaginal orifice closure. Their coordinated contraction relies on both the endurance of slow-twitch muscle fibers and the explosive strength of fast-twitch muscle fibers. When there is an imbalance in muscle force distribution—such as a decline in slow-twitch fiber endurance or a weakening of fast-twitch fiber contraction—the surrounding musculature fails to coordinate effectively, resulting in incomplete closure^38^. Specifically, imbalances in coronal plane muscle forces (e.g., reduced tension in the pubovaginalis muscle) prevent close adherence between the anterior and posterior walls of the vagina. Concurrently, insufficiencies in sagittal plane muscle forces (notably diminished longitudinal support from the levator ani complex) lead to inadequate upward pulling at the lower end of the vaginal orifice, culminating in a “funnel-shaped” relaxation pattern. Traditional PFM-assessment methods can partially assist to detect the vaginal-orifice relaxation caused by insufficient synthetical PFM strength of the whole vagina muscle. In contrast, our 3D accurate PFM-distribution detecting method not only solves the problem of evaluating the UVO degree, but also facilitates to find the root cause of the precise relaxation foci of the vaginal wall and orifice. Therefore, the high-precision PFM-strength resolution combined with multi-channel 3D spatial resolution allows for the precise identification of local PFM-strength defects, providing quantitative basis for the localization diagnosis of PFD.

#### (3) PFM-strength characteristics of postpartum SUI subgroup

Female SUI is closely associated with abnormal PFM strength. Traditional cognition of SUI shows that the contraction EMG potentials for women with SUI are usually weaker than those of healthy women^39^. This superficial connection is not enough to make pelvic-floor EMG become a SUI-identifying method. In this work, our directly accurate multi-channel PFM-strength measuring method was used to seek the relationship between PFM-strength characteristics and SUI symptom for postpartum women. Detailed PFM-strength distributions of 14 SUI subjects have been systematically measured and analyzed. The 3D PFM-strength distributions could provide more physiological information to assist doctors’ principle analysis and classification diagnosis. Significantly, our 3D high-precision PFM-strength monitoring method is conducive to construct “PFM strength-anatomy-symptom” correlation model, providing basis for the classification diagnosis of PFD. Several typical PFM-strength distribution characteristics of postpartum SUI subjects have been found, including 1) abnormal weak PFM strength of the whole vagina muscle (Fig. 5a), 2) UVO-related SUI with extremely weak PFM-strength distributions in the distal region (Fig. 5b), and 3) weaker PFM-strength of slow-twitch muscle fibers than that of fast-twitch muscle fibers (Fig. 5c). Figure 5d shows the 3D PFM-strength distributions in four directions of vagina: anterior, right, posterior, and left, which are corresponding to a general PFM-strength weakening subject and a UVO-related SUI subject, respectively. The typical multi-channel PFM strength distributions for individual subjects distinguished by the three classification are shown in Supplementary Figs. 3a−c. Abnormal weak PFM strength of the whole vagina muscle could lead to downward displacement of the bladder and urethra, which is the anatomical pathological basis of SUI. UVO-related SUI is generally caused by the closure mechanism disorder of the urethra, which is consistent with the clinical reality of SUI. Weak slow-twitch muscle strength is consistent with pelvic floor support disorder which could result in bladder displacement. These three typical PFM-strength distribution characteristics as statistically analyzed in Figs. 5a−c, which fully reveal the pathogenesis of SUI, have not been found in currently existing PFM evaluating methods^15–25^.

**Fig. 5.**
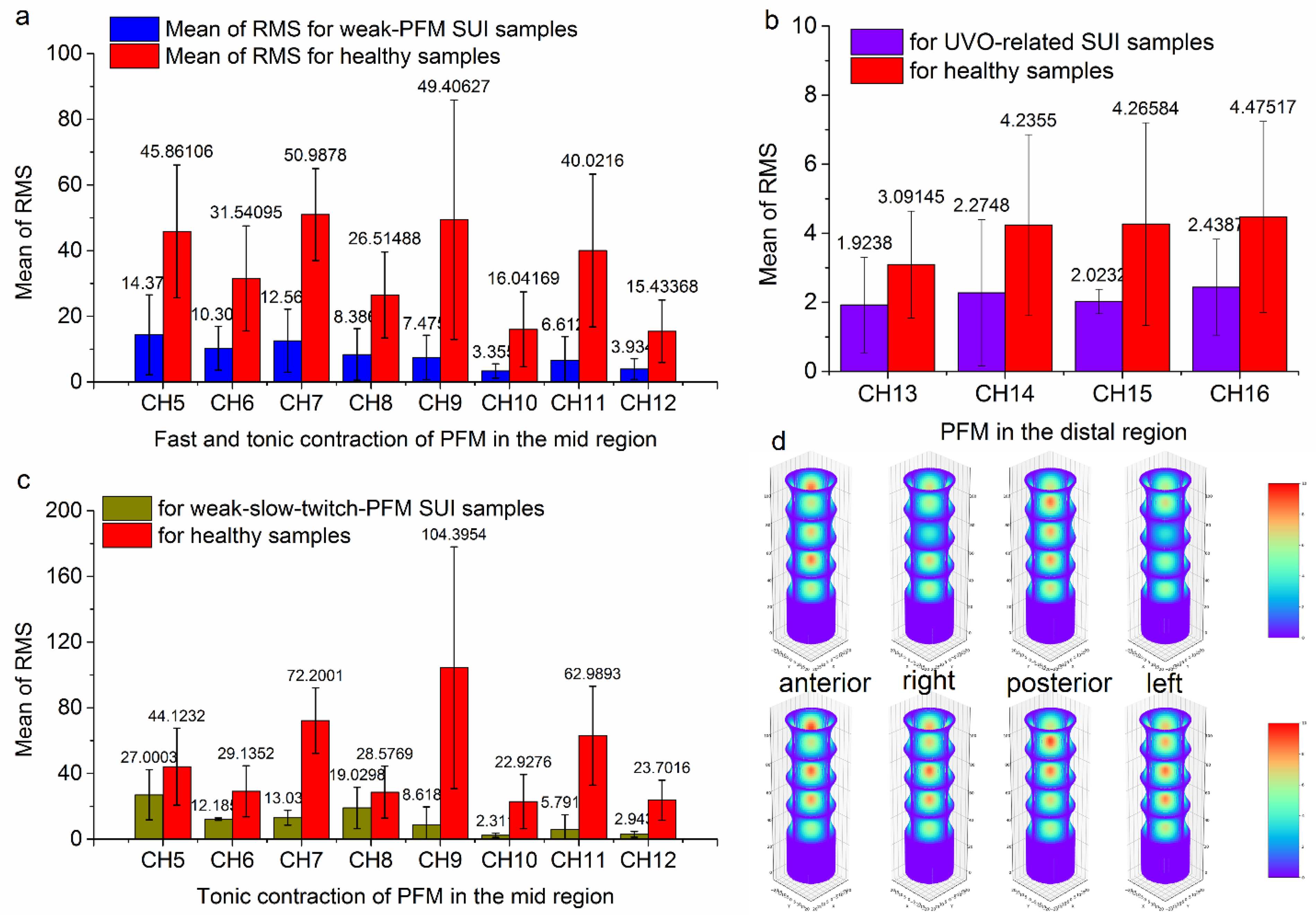
Comparison of PFM-strength characteristics for classified postpartum SUI subgroup and healthy group. **a** RMS of local PFM strength at each channel in the mid region for the general PFM weakening type SUI subgroup compared with that of healthy group. **b** RMS of local PFM strength at each channel in the distal region for UVO-related SUI subgroup compared with that of healthy group. **c** RMS of local PFM strength at each channel in the mid region for the slow-twitch PFM weakening type SUI subgroup compared with that of healthy group. **d** The comparison of 3D PFM-strength-distribution in four directions of vagina: anterior, right, posterior, and left for a general PFM-strength weakening subject (shown in upper line) and a UVO-related SUI subject (shown in bottom line), respectively.

The SUI-related muscles mainly include the urethral sphincter, levator ani muscles, transverse perineal muscle, bulbospongiosus muscle, and external anal sphincter. These muscles play a crucial role in maintaining urethral closure and urinary control through coordinated action^40^. The urethral sphincter encircles the urethra in an omega shape and maintains closure by contracting and compressing the urethra. The transverse perineal muscle plays an indirect yet crucial role in supporting the pelvic floor organs and influencing urethral closure through its involvement in the formation of the perineal central tendon. The bulbospongiosus muscle contributes to maintain urethral closure by contracting and narrowing the vaginal canal. Any abnormality in muscle strength may compromise this function. Furthermore, the external anal sphincter ensures pelvic floor stability through coordinated actions with other pelvic floor muscles. Diminished muscle strength can adversely affect urethral closure, potentially leading to SUI. Slow-twitch muscle fibers provide continuous tension to sustain resting pressure around the urethra, while fast-twitch muscle fibers contract rapidly when abdominal pressure increases, further enhancing compression of the urethra. Damage to these muscle fibers can impair effective closure of the urethra, resulting in SUI disease. The levator ani muscles—comprising the iliococcygeus, pubococcygeus, and puborectalis—offer elastic support for the urethra through sustained contraction of type I (slow-twitch) muscle fibers and maintain the urethra at the normal anatomic place. Additionally, they facilitate rapid closure of the urethra during episodes of increased abdominal pressure via type II (fast-twitch) muscle fibers. Weakened muscular strength or dysfunction may lead to inadequate support for the urethra and elevate the risk of SUI. Significantly, compared with traditional electrical stimulation therapy that only have effect on PFM in the midpiece region without distinguishing specific local area, our accurate PFM-strength distributions measuring method also offer opportunities to find the location of lesions and simultaneously playing an important role in precise diagnosis and treatment. In addition, our accurate 3D PFM-strength detecting method can also quantify the changes in local PFM strength before and after treatment, as shown in Supplementary Fig. 3d, providing objective indicators for the curative effect.

## DISCUSSION

This work proposed a direct method for accurately detecting 3D PFM-strength distributions. The resulting accurate PFM state and functional assessment with high sensitivity and spatial resolution could provide biomechanical foundation for clinical early physiotherapy diagnostic and precise treatment of PFD. The high sensitivity with PFM-strength resolution of 1E-4 cmH_2_O permits observations of abnormally weak PFM strength state, allowing the identification of subclinical-state functional abnormalities before clinical symptoms appearing. As a demonstration, subclinical PFM-strength abnormalities in the postpartum AWO population have been quantified, offering opportunities for early warning and primary prevention of PFD during the asymptomatic period. More importantly, the combination of high sensitivity with multi-channel spatial resolution allows for the identification of local PFM-strength defects, providing quantitative basis for the localization diagnosis of PFM defects. This has been well demonstrated by the clinical trials for postpartum UVO and SUI population. In addition, compared with traditional PFM-evaluating methods^15–25^, there are three-aspect advantages from our developed PFM-strength measuring system. First, our novel designed vaginal probe exhibited additional advance in physical comfortability, patients-friendliness universality, and stability without motion artifact. Second, our system could release the requirements of subjective judgment and personal experience of clinicians by remarkably improving the accuracy and reliability of PFM-strength assessments while minimizing biases stemming from variations in physician experience—thereby promoting standardized diagnosis and treatment across diverse medical settings. Specifically, the portability is conducive to quickly deploy our PFM-strength dynamic measuring system in the outpatient department, operating room, or even community settings, enhancing and expanding the operability of PFD-related screening. The resulting objective and quantitative 3D PFM-strength distributions would offer revolutionary support for clinicians to explore the underlying pathophysiological mechanisms of PFD and devise more targeted intervention strategies for different PFD symptoms.

Looking forward to future practical clinical application, this study still has at least two-aspect limitations. One is about the selection of sample group. We enrolled 51 postpartum women with early PFD symptoms and 29 healthy women to participate in this prospective study. For the postpartum group, we selected subjects after CS or VD within 6-12 weeks to deeply participate in this study. The specific selection of the postpartum sample group is conducive to conduct targeted analysis for the early characteristics of PFD-related disease, since it has been well demonstrated that pregnancy and childbirth are etiological factors in PFD pathogenesis^41–44^. Nevertheless, the narrow postpartum time-frame selection would limit the expanding applications of the innovative method. In fact, the accurate detection of 3D PFM-strength distributions developed in this work, which offers obvious advantages in terms of high accuracy, high sensitivity, and high spatial resolution, would provide revolutionary support for clinicians to perform early screening and devise targeted intervention strategies for different PFD symptoms in various female groups. It is well known that postpartum and post-menopausal elderly female groups are the two major populations that are prone to PFD. In the next stage, we will focus on the accurate evaluation of PFM-strength distributions in post-menopausal elderly women to enable expanded early diagnosis and precise treatment of PFD. The other is about the lack of referencing standard. Different from traditional PFM-evaluating methods^15–25^, our direct method for accurately detecting 3D PFM-strength distributions based on a novel designed PMVP with multi-channel EGPSs lacks effective references of existing similar methods or standards. To reveal the feasibility and validity of our method, clinical PFM strength acquisition comparisons were conducted using our novel multi-channel PFM-strength measuring method and commercial pelvic-floor EMG method (MLD B4Plus, Nanjing Mai Land Medical Technology Co., Ltd., China). To further confirm the validity of our 3D PFM-strength distribution detecting method, gold-standard ultrasound imaging comparison has also been conducted. However, to fully take advantages of high PFM-strength sensitivity and 3D spatial resolution that other methods do not have in practical clinical applications, the standardization is highly required by establishing new standards for our innovative method.

Currently clinical trials show ability of our method in linking various early PFD symptoms (e.g. AWO, UVO and SUI) with certain abnormal PFM-strength distribution characteristics, providing a visual biomechanical foundation for clinical early diagnosis and the classification diagnosis of PFD. From long-term application perspective, the value of the 3D PFM-strength distribution detecting approach is far more than enhancing diagnostic capabilities. On the one hand, individualized precision rehabilitation programs with monitoring treatment effects and improving patient prognosis could be expected. For example, targeted endurance training can be formulated for patients with slow muscle fiber weakness such as AWO conditions shown in Fig. 3. Targeted Kegel exercises can be performed for patients with weak PFM strength in the perineal area of the external vagina orifice such as UVO conditions shown in Fig. 4. O<u>ur</u> accurate 3D PFM-strength detecting method can quantify the changes in local PFM strength before and after treatment, as demonstrated in Supplementary Fig. 3d, providing objective indicators for curative effect. The rapid feedback mechanism regarding treatment responses facilitates timely adjustments to therapeutic plans, ultimately shortening rehabilitation periods while optimizing resource utilization and enhancing patient satisfaction. On the other hand, the novel design of PMVP leaves room for future integration of multi-channel synchronous electrical stimulation functions. Multi-channel stimulated electrodes will be prepared on the surface of multi-contacts of PMVP, offering customized precise electrical stimulation therapy according to corresponding PFM-strength distributions for individuals. In this way, a new concept of 3D PFM detection and treatment will be expected to promote the paradigm revolution in PFD-related clinical early diagnosis and precise treatments. From a health economics perspective, this technology effectively reduces the demand for invasive treatments and costly surgeries in the later stages of disease progression by enabling early accurate diagnosis along with precise intervention. Furthermore, it alleviates unnecessary referrals and diagnostic tests, significantly enhancing the efficiency of medical resource allocation.

In conclusion, this study demonstrated a direct method for accurately detecting 3D PFM-strength distributions by developing a portable multi-channel PFM-pressure dynamic measuring system. Compared with currently existing PFM-evaluating methods, this method offers significant advantages in term of high PFM-strength sensitivity and 3D spatial resolution, enabling precise visualization and quantitative analysis of PFM functions. The clinical application value has been exhibited in the following three aspects: (1) The diagnostic window would be significantly advanced due to the ability to identify subclinical PFM-strength abnormalities, e.g. AWO, achieving early warning. (2) PFM-strength evaluation has been extended from one-dimensional comprehensive force to three-dimensional PFM-strength distribution, providing quantitative basis for the localization diagnosis and the classification diagnosis of PFD. (3) Rehabilitation training could be upgraded from universal guidance to personalized navigation, promoting precision rehabilitation. Despite limitations from samples and lacking referencing standard, the long-term developing value of the 3D PFM-strength detecting approach is still worthy looking forward. On the one hand, individualized precision rehabilitation programs with monitoring treatment effects and improving patient prognosis could be expected due to the scientific foundation offered by the individualized accurate 3D PFM-strength distributions. On the other hand, the novel design of PMVP leaves room for future integration of multi-channel synchronous electrical stimulation treatments, offering customized precise electrical stimulation therapy according to corresponding PFM-strength distributions for individuals. Combining additional advantages of the portable measuring system in terms of physical comfortability, patients-friendliness universality, and stability without motion artifact, new paradigm with personalized precise electrical stimulation and rapid feedback mechanism would be stimulated. With ongoing optimization of the technology coupled with an expansion in its clinical applications, this method will significantly improve both diagnosis and treatment quality for PFD population, ultimately enhancing their life quality while advancing the entire field of pelvic floor medicine.

## Methods

### Ethics statement

This study was approved by the Research Medical Ethics Committee of the West China Second University Hospital, Sichuan University (IRB approval number: 2023317). Written informed consent was obtained from all participants involved in the study. Personal identifiers were completely removed and the data were analyzed anonymously. This study was conducted according to the ethical standards recommended by the 1964 Declaration of Helsinki and its later amendments.

### Study population

We have calculated the required sample size with power analysis before carrying out clinical trials. A sample size of 29 produces a two-sided 95% confidence interval with a width equal to 0.2 when the sample proportion is 0.95. We enrolled 51 postpartum women with early PFD symptoms and 29 healthy women to participate in this prospective study. Significantly, these 29 healthy subjects are without any PFD symptoms, without postpartum restriction, and just with an interested in using the novel PFM-measuring system. They were used as reference group to conduct contrastive analysis for the early PFD-related PFM characteristics for the postpartum group. These 51 postpartum women, who aged between 20 and 40 years after cesarean section (CS) or vaginal delivery (VD) within 4-12 weeks, were deeply participated in this study. The specific selection of the postpartum sample group is conducive to conduct targeted analysis for the early characteristics of PFD-related disease, since it has been well demonstrated that pregnancy and childbirth are etiological factors in PFD pathogenesis^41–44^. The underlying mechanism involves structural alterations and trauma to pelvic floor anatomy during gestational and parturition processes^45–47^. Exploring complex 3D PFM strength of postpartum participants is crucial to analyze the multifactorial damage to the vaginal wall with surrounding muscles and fascia, facilitating to PFD diagnosis and treatment at an early stage. The sample size of the subgroup AWO, UVO and SUI were 22, 15, and 14, respectively.

### PFM measurements

We performed a cohort study at West China Second University Hospital of Sichuan University to explore complex 3D PFM-strength distributions. A comprehensive pelvic floor examination has been performed for all subjects, including sequential vaginal digital palpation, intravaginal EMG, and our real-time dynamic multi-channel PFM pressure measurement. The Glazer protocols was selected as the standard assessment actions, which consists of a rest, a rapid contraction, and an endurance contraction^48^. The rapid contraction was used to assess functions of the fast-twitch muscle fibers, while the endurance contraction was used to assess functions of the slow-twitch muscle fibers. The subjects were instructed to empty their bladders, perform vaginal digital palpation, and then experience a clinical standard single-channel EMG test. During vaginal digital palpation, subjects were asked to lay in supine position with 45 degree of hip and knee relaxed after voiding urine. Distal phalanx of index and middle finger of clinical trial doctor was placed in distal vagina, and subjects were asked to squeeze and lift the pelvic floor. Then, a rough and overall PFM-strength state was evaluated according to Modified Oxford Grading Scale. During intravaginal EMG measurements, subjects were also asked to lay in supine position with 45 degree of hip and knee relaxed after voiding urine. The EMG vaginal probe was cleaned and disinfected in advance. The EMG vaginal probe was placed in the vagina of the subjects. A pair of reference electrodes were placed on the abdomen bony area. Another pair of electrodes were placed in the region of abdominal rectus muscles to monitor crosstalk of contraction of the abdominal muscles. A two-step procedure according to the Glazer protocols was conducted to indirectly access the strength of Type I and Type II muscle fibers in pelvic floor^49^. EMG signal during five-times fast contraction and relax was used to evaluate Type I muscle fibers, which contribute to pelvic movement and urinary control and are mainly present in the superficial perineal muscles. EMG signal during sustained contraction for more than 5 seconds was used to reveal Type II muscle fibers, which support pelvic and abdominal organs and are primarily found in the levator ani muscle.

During multi-channel 3D PFM-strength measurement, the same standardized protocol according to the Glazer protocols was utilized to ensure reproducibility and accuracy across different clinical settings. The measurement protocol consisted of three distinct phases: resting state assessment, dynamic contraction series, and endurance evaluation. First, subjects were also asked to lay in supine position with 45 degree of hip and knee relaxed after voiding urine. The PMVP that was cleaned and disinfected in advance was covered with unlubricated condoms, coated with vaginal lubricant, and then placed into vagina of subject. Subjects were asked to relax by maintaining normal breathing and refraining from any voluntary contraction of the pelvic floor. The PFM measuring system conducted an automated baseline recording of the resting pressures across all 17 sensor nodes, thereby generating a comprehensive pressure profile map of the vaginal canal at rest. Then, the real-time dynamic multi-channel PFM-strength measurement was performed based on two-step procedure: 1) fast contraction and relax for five times; 2) tonic contraction for five times. Prior to the examination, participants received standardized instructions on correct PFM contraction technique, emphasizing the importance of isolating the PFM without activating accessory muscles (abdominals, gluteals, and adductors). Specifically, it is crucial to minimize the involvement of abdominal muscles during PFM contractions. Our approach can monitor abdominal pressure that confused into PFM strength by the design of domal contact with EGPS, ensuring more accurate data regarding pelvic floor muscle activity. Comparable to clinical used EMG testing method, the functions of Type I and Type II muscle fibers could also be evaluated by using the multi-channel PFM pressure results during fast contraction and tonic contraction process. This is critical to confirm the validation of our novel PFM-strength measuring method. More importantly, accurate multi-channel PFM-strength data can be directly obtained to construct PFM-strength distributions derived from various local pelvic floor muscles (e.g. vaginal sphincter, urethral sphincter, puborectalis, pubococcygeus muscles, and lliococcygeus muscles) at their specific locations. The 3D mapping of PFM-strength distributions will provide a detailed partitioned PFM diagram, establishing a reliable theoretical foundation for subsequent diagnosis and treatment of PFD.

### Statistical analysis

The sample size was analyzed by using PASS 2021 (Version 21.0.3). PFM strength data in each channel was expressed as RMS. The consistency and difference for each channel between different samples were expressed as mean of RMS with standard deviation (SD). A *P* value < 0.05 was considered statistically significant and analyzed by one-way ANOVA. All PFM-strength distributions’ data statistics were analyzed by Origin (Version 9.0).

## Data Availability

All data produced in the present study are available upon reasonable request to the authors.

## Acknowledgements

This work was supported by the National Key Research and Development Program of China (2021YFC2009100), the Science and Technology Department of Sichuan Province Project (2023YFS0024), and the Sichuan Provincial Health Commission Project (21ZD002). The funding body played no role in the study design, data collection, analysis and interpretation, or manuscript writing.

## Author contributions

Conceptualization: X.N., Y.L., N.L. and T.W.; Methodology: H.W., N.L. and Y.L.; Formal Analysis: N.L., T.W. and H.W.; Investigation: T.W., Y.Z., X.Y., Q.W., D.W., R.L. and L.D.; Visualization: H.W., N.L., Y.Z. and D.W.; Funding Acquisition: X.N. and Y.L.; Project Administration: T.W., X.N. and Y.L.; Supervision: T.W., Y.L. and X.N.; Writing – Original Draft: N.L., Y.L., T.W. and X.N.; Editing: N.L., T.W., Y.L. and X.N.

## Competing interests

All authors declare no financial or non-financial competing interests.

## Data and materials availability

Raw data are available upon request.

## Notes

### Competing Interest Statement

The authors have declared no competing interest.

### Funding Statement

This work was funded by the National Key Research and Development Program of China (2021YFC2009100), the Science and Technology Department of Sichuan Province Project (2023YFS0024), and the Sichuan Provincial Health Commission Project (21ZD002).

### Author Declarations

The Research Medical Ethics Committee of the West China Second University Hospital in Sichuan University gave ethical approval for this work.

## References

1. Dubik, J., Alperin, M. & De Vita, R. The biomechanics of the vagina: a complete review of incomplete data. npj Women’s Health 3, 4 (2025).

2. Duran, P. et al. Proregenerative extracellular matrix hydrogel mitigates pathological alterations of pelvic skeletal muscles after birth injury. Sci. Transl. Med. 15, eabj3138 (2023).

3. Dietz, H. P., Shek, C. & Clarke, B. Biometry of the pubovisceral muscle and levator hiatus by three - dimensional pelvic floor ultrasound. Ultrasound in Obstet. & Gyne. 25, 580–585 (2005).

4. DeLancey, J. O. L. The anatomy of the pelvicfloor. Curr. Opin. Obstet. Gynecol. 6, 313–316 (1994).

5. Boscolo Sesillo, F., et al. Effect of lactation on postpartum pelvic floor muscle regeneration in preclinical model. npj Women’s Health 3, 33 (2025).

6. DeLancey, J. O. L. et al. Pelvic floor injury during vaginal birth is lifealtering and preventable: what can we do about it?. Am. J. Obstet. Gynecol. 230, 279–294 e272 (2024).

7. Blomquist, J. L., Carroll, M., Munoz, A. & Handa, V. L. Pelvic floor muscle strength and the incidence of pelvic floor disorders after vaginal and cesarean delivery. Am. J. Obstet. Gynecol. 222, 62.e1–62.e8 (2020).

8. Hallock, J. L. & Handa, V. L. The epidemiology of pelvic floor disorders and childbirth: An update. Obstet. Gynecol. Clin. North Am. 43, 1–13 (2016).

9. Wu, J. M., et al. Prevalence and trends of symptomatic pelvic floor disorders in U.S. women. Obstet. & Gyne. 123, 141–148 (2014).

10. Xia, W., et al. Automatic plane of minimal hiatal dimensions extraction from 3D female pelvic floor ultrasound. IEEE Trans. Med. Imaging 41, 3873–3883 (2022).

11. Peinado-Molina, R. A., Hernandez-Martinez, A., Martinez-Vazquez, S., Rodriguez-Almagro, J. & Martinez-Galiano, J. M. Pelvic floor dysfunction: prevalence and associated factors. BMC Public Health 23, 2005 (2023).

12. Shen, L., Yang, J., Bai, X. & Sun, Z. Analysis of the current status of pelvic floor dysfunction in urban women in Xi’an City. Ann. Palliat. Med. 9, 979–984 (2020).

13. Handa, V. L., Roem, J., Blomquist, J. L., Dietz, H. P. & Muñoz, A. Pelvic organ prolapse as a function of levator ani avulsion, hiatus size, and strength. Obstet. & Gynecol. Surv. 74, 588–590 (2019).

14. Yaşar, L., Telci, S. O., Doğan, K., Kaya, E. & Ekin, M. Predictive role of measurement of pelvic floor muscle thickness with static MRI in stress and mixed urinary incontinence. Int Urogynecol J 30, 271–277 (2019).

15. DaSilva, J. B., De Oliveira Sato, T., Rocha, A. P. R. & Driusso, P. Inter- and intrarater reliability of unidigital and bidigital vaginal palpation to evaluation of maximal voluntary contraction of pelvic floor muscles considering risk factors and dysfunctions. Neurourol. Urodynam. 40, 348–357 (2021).

16. Deegan, E. G., Stothers, L., Kavanagh, A. & Macnab, A. J. Quantification of pelvic floor muscle strength in female urinary incontinence: A systematic review and comparison of contemporary methodologies. Neurourol. Urodynam. 37, 33–45 (2018).

17. Bø, K. & Finckenhagen, H. B. Vaginal palpation of pelvic floor muscle strength: inter-test reproducibility and comparison between palpation and vaginal squeeze pressure. Acta Obstet Gynecol Scand 80, 883–883 (2001).

18. Chamochumbi, C. C. M., Nunes, F. R. Guirro, R. R. J. & Guirro, E. C. O. Comparison of active and passive forces of the pelvic floor muscles in women with and without stress urinary incontinence. Rev. bras. fisioter. 16, 314–319 (2012).

19. Morin, M., Gravel, D., Bourbonnais, D., Dumoulin, C. & Ouellet, S. Reliability of dynamometric passive properties of the pelvic floor muscles in postmenopausal women with stress urinary incontinence. Neurourol. Urodynam. 27, 819–825 (2008).

20. Dumoulin, C., Gravel, D., Bourbonnais, D., Lemieux, M. C. & Morin, M. Reliability of dynamometric measurements of the pelvic floor musculature. Neurourol. Urodynam. 23, 134–142 (2004).

21. Hundley, A. F., Wu, J. M. & Visco, A. G. A comparison of perineometer to brink score for assessment of pelvic floor muscle strength. AM J Obstet. Gynecol. 192, 1583–1591 (2005).

22. Da Silva Borin, L. C. M., Nunes, F. R. & De Oliveira Guirro, E. C. Assessment of pelvic floor muscle pressure in female athletes. PM&R 5, 189–193 (2013).

23. Enck, P. & Vodušek, D. B. Electromyography of pelvic floor muscles. Journal of Electromyography and Kinesiology 16, 568–577 (2006).

24. Grape, H. H., Dedering, A. & Jonasson, A. F. Retest reliability of surface electromyography on the pelvic floor muscles. Neurourology and Urodynamics 28, 395–399 (2009).

25. Flury, N., Koenig, I. & Radlinger, L. Crosstalk considerations in studies evaluating pelvic floor muscles using surface electromyography in women: a scoping review. Arch. Gynecol. Obstet. 295, 799–809 (2017).

26. Brink, C. A., Wells, T. J., Sampselle, C. M., Taillie, E. R. & Mayer, R. A digital test for pelvic muscle strength in women with urinary incontinence. Nursing Research 43, 352–356 (1994).

27. Keshwani, N. & McLean, L. State of the art review: Intravaginal probes for recording electromyography from the pelvic floor muscles. Neurourol. Urodynam. 34, 104–112 (2015).

28. Wang, S., et al. Physiology-based stretchable electronics design method for accurate surface electromyography evaluation. Adv. Sci. 8, 2004987 (2021).

29. Huntington, A., Donaldson, K. & De Vita, R. Contractile properties of vaginal tissue. Journal of Biomechanical Engineering 142, 080801 (2020).

30. Skoczylas, L. C. et al. Regional differences in rat vaginal smooth muscle contractility and morphology. Reprod. Sci. 20, 382–390 (2013).

31. Giraldi, A. et al. Morphological and functional characterization of a rat vaginal smooth muscle sphincter. Int J Impot Res 14, 271–282 (2002).

32. Clark, G. L. et al. Smooth muscle regional contribution to vaginal wall function. Interface Focus. 9, 20190025 (2019).

33. Oh, S.-J., Hong, S. K., Kim, S. W. & Paick, J.-S. Histological and functional aspects of different regions of the rabbit vagina. Int J Impot Res 15, 142–150 (2003).

34. Basha, M. et al. Regional differences in myosin heavy chain isoform expression and maximal shortening velocity of the rat vaginal wall smooth muscle. *American Journal of Physiology-Regulatory*, Integrative and Comparative Physiology 291, R1076–R1084 (2006).

35. Śmietański, M., Śmietańska, I. A. & Zamkowski, M. Post-partum abdominal wall insufficiency syndrome (PPAWIS): lessons learned from a single surgeon’s experience based on 200 cases. BMC Surg 22, 305 (2022).

36. Gao, Q., et al. Pelvic floor dysfunction in postpartum women: A cross-sectional study. PLoS ONE 19, e0308563 (2024).

37. Urbankova, I. et al. First delivery and ovariectomy affect biomechanical and structural properties of the vagina in the ovine model. Int Urogynecol J 30, 455–464 (2019).

38. Bø, K., et al. Recovery of pelvic floor muscle strength and endurance 6 and 12 months postpartum in primiparous women—a prospective cohort study. Int Urogynecol J 33, 3455–3464 (2022).

39. Yang, X., et al. Comparisons of electromyography and digital palpation measurement of elvic floor muscle strength in postpartum women with stress urinary incontinence and asymptomatic parturients: A cross-sectional study. Gynecol Obstet Invest 84, 599–605 (2019).

40. Yang, X., et al. The Anatomical Pathogenesis of Stress Urinary Incontinence in Women. Medicina 59, 5 (2022).

41. Mothes, A. R., Radosa, M. P., Altendorf-Hofmann, A. & Runnebaum, I. B. Risk index for pelvic organ prolapse based on established individual risk factors. Arch Gynecol Obstet 293, 617–624 (2016).

42. Romeikienė, K. E. & Bartkevičienė, D. Pelvic-floor dysfunction prevention in prepartum and postpartum periods. Medicina 57, 387 (2021).

43. Van Geelen, H., Ostergard, D. & Sand, P. A review of the impact of pregnancy and childbirth on pelvic floor function as assessed by objective measurement techniques. Int Urogynecol J 29, 327–338 (2018).

44. He, R., et al. The effect of pelvic floor muscle training and perineal massage in late pregnancy on postpartum pelvic floor function in nulliparas: A randomised controlled clinical trial. Complement. Ther. Med. 77, 102982 (2023).

45. Aasheim, V., Nilsen, A. B. V., Reinar, L. M. & Lukasse, M. Perineal techniques during the second stage of labour for reducing perineal trauma. Cochrane Database of Systematic Reviews 6, CD006672 (2017).

46. Woodley, S. J., et al. Pelvic floor muscle training for preventing and treating urinary and faecal incontinence in antenatal and postnatal women. Cochrane Database of Systematic Reviews 5, CD007471 (2020).

47. Blomquist, J. L., Muñoz, A., Carroll, M. & Handa, V. L. Association of delivery mode with pelvic floor disorders after childbirth. JAMA 320, 2438–2447 (2018).

48. Romanzi, L. J., Polaneczky, M. & Glazer, H. I. Simple test of pelvic muscle contraction during pelvic examination: Correlation to surface electromyography. Neurourol. Urodynam. 18, 603–612 (1999).

49. Aljuraifani, R., Stafford, R. E., Hall, L. M., Van Den Hoorn, W. & Hodges, P. W. Task-specific differences in respiration-related activation of deep and superficial pelvic floor muscles. *J.Appl*. Phys. 126, 1343–1351 (2019).

