## Supplementary figures and images for "Accurate detection of 3D pelvic-floor-muscle strength for reforming clinical diagnosis and treatment of women’s pelvic floor dysfunction"

### Supplementary Figure 1

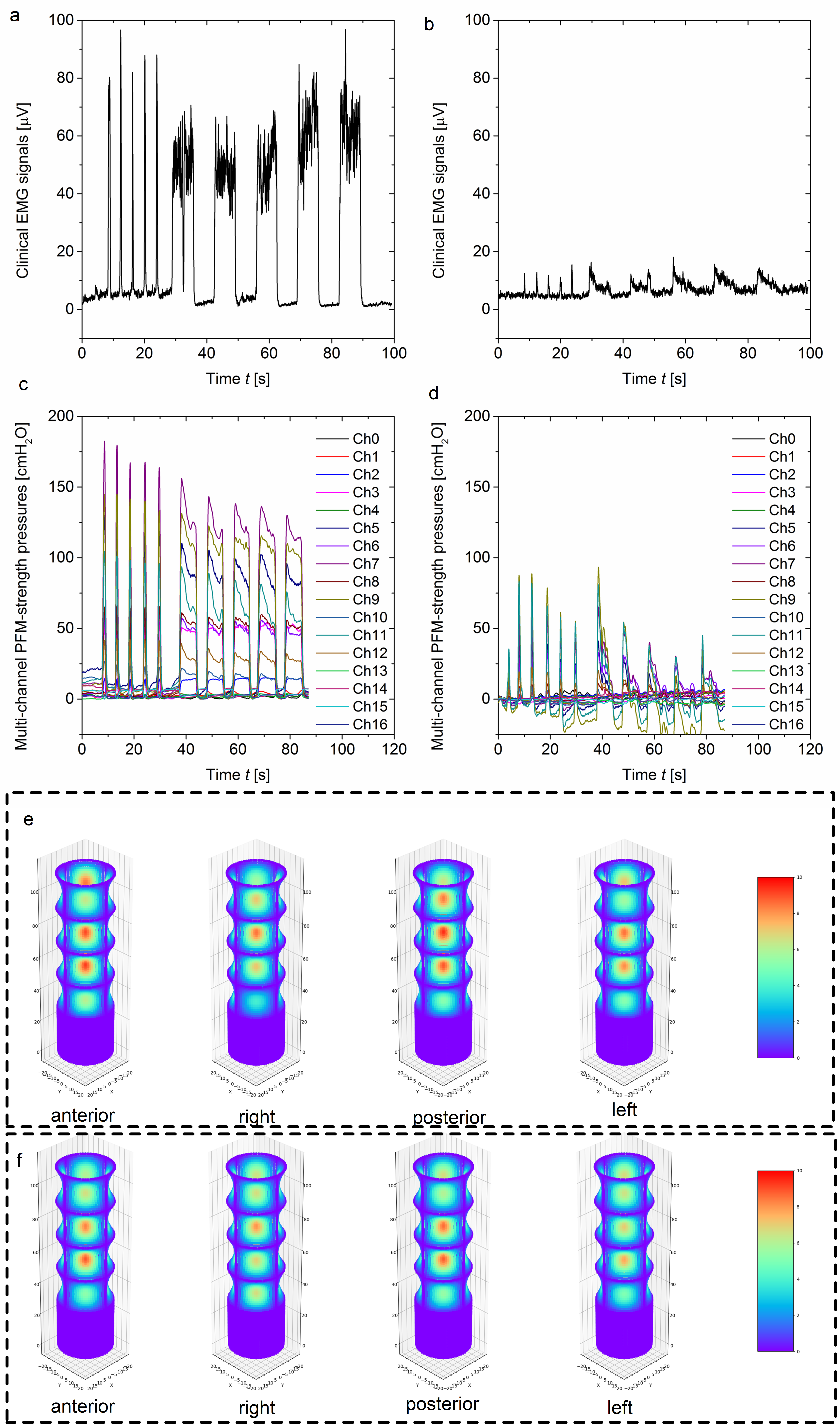

### Supplementary Figure 2

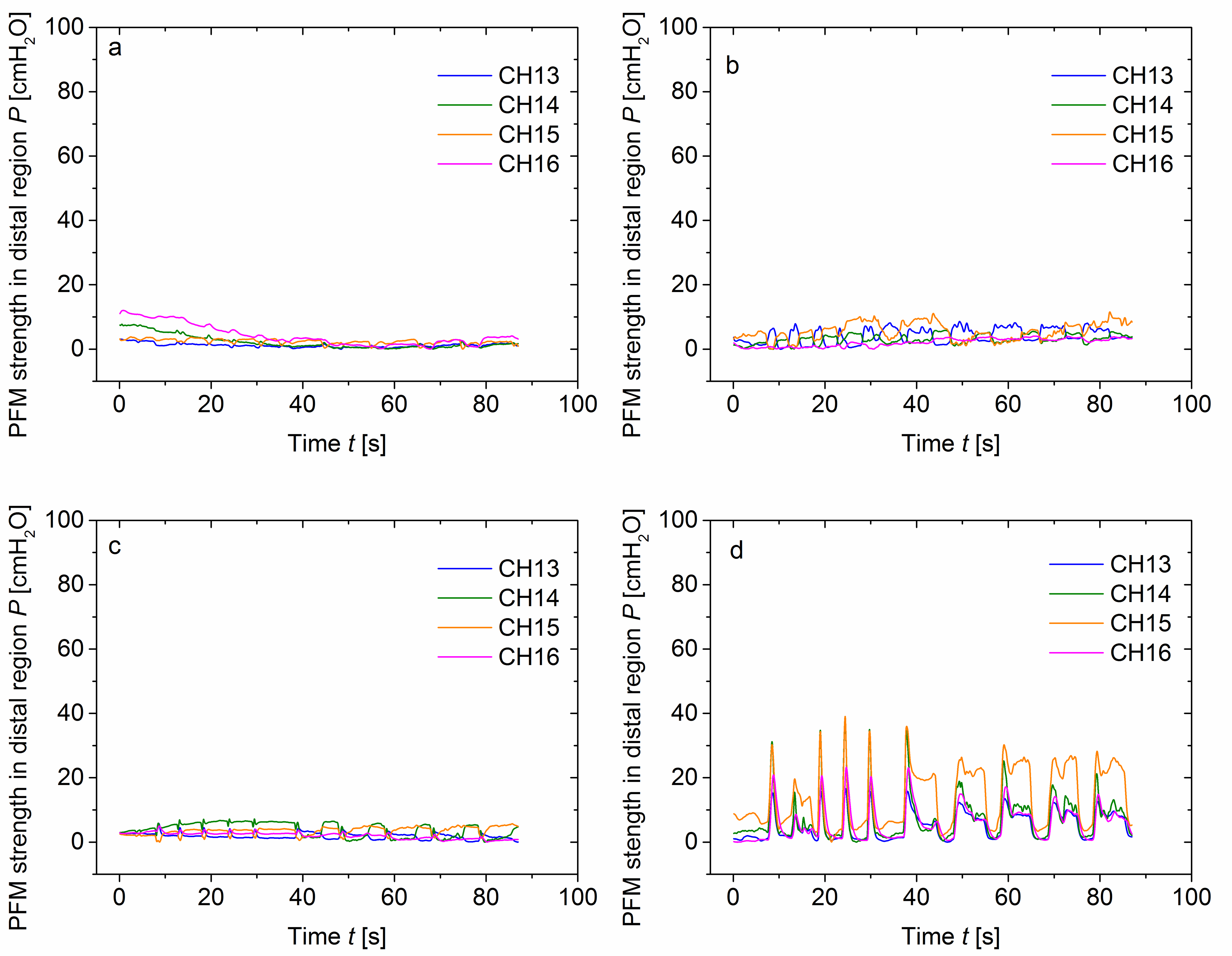

### Supplementary Figure 3

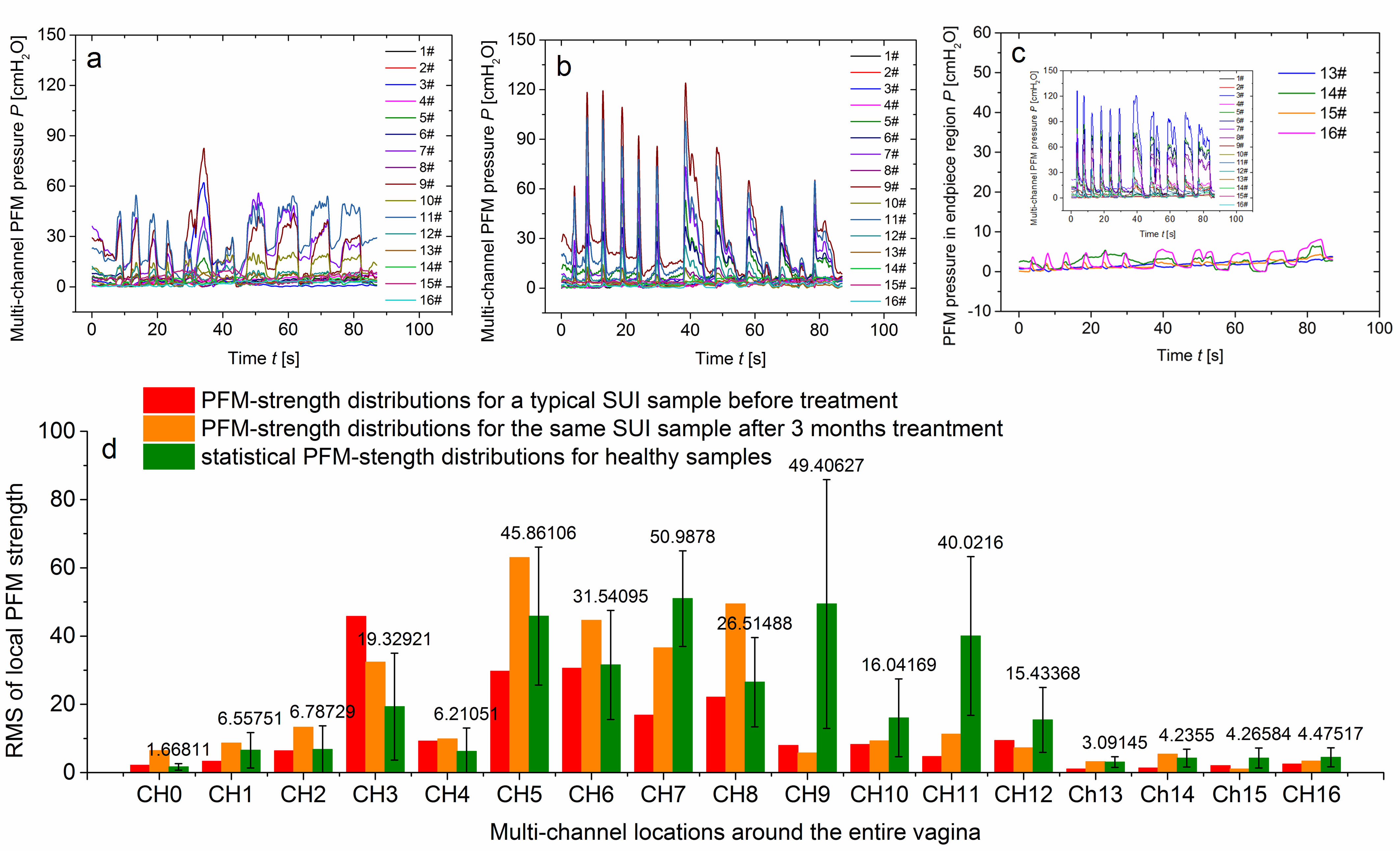
